# Individualizing deep dynamic models for psychological resilience data

**DOI:** 10.1101/2020.08.18.20177113

**Authors:** Göran Köber, Shakoor Pooseh, Haakon Engen, Andrea Chmitorz, Miriam Kampa, Anita Schick, Alexandra Sebastian, Oliver Tüscher, Michèle Wessa, Kenneth S.L. Yuen, Henrik Walter, Raffael Kalisch, Jens Timmer, Harald Binder

**Affiliations:** Institute of Medical Biometry and Statistics (IMBI), Faculty of Medicine and Medical Center, University of Freiburg, 79104, Germany; Freiburg Center of Data Analysis and Modelling (FDM), University of Freiburg, Freiburg, 79104, Germany; Institute of Physics, University of Freiburg, 79104, Germany; Neuroimaging Center (NIC), Focus Program Translational Neuroscience (FTN), Johannes Gutenberg University Medical Center, Mainz, 55131, Germany; Leibniz Institute for Resilience Research (LIR), Mainz, 55122, Germany; Department of Psychiatry and Psychotherapy, Johannes Gutenberg University Medical Center, Mainz, 55131, Germany; Faculty of Social Work, Health and Nusing, University of Applied Sciences Esslingen, Esslingen, 73728, Germany; Department of Clinical Psychology, University of Siegen, 57076, Germany; Bender Institute of Neuroimaging (BION), Department of Psychology, Justus Liebig University, Gießen, 35394, Germany; Department of Public Mental Health, Central Institute of Mental Health, Medical Faculty Mannheim, Heidelberg University, Germany; Department of Clinical Psychology and Neuropsychology, Institute of Psychology, Johannes Gutenberg University, Mainz, 55131, Germany; Research Division of Mind and Brain, Charité–Universitätsmedizin Berlin, Corporate Member of Freie Universität Berlin, Humboldt-Universität zu Berlin, and Berlin Institute of Health, Germany; Berlin School of Mind and Brain, Humboldt-Universität zu Berlin, Germany; CIBSS—Centre for Integrative Biological Signaling Studies, University of Freiburg, 79104, Germany

## Abstract

Deep learning approaches can uncover complex patterns in data. In particular, variational autoencoders (VAEs) achieve this by a non-linear mapping of data into a low-dimensional latent space. Motivated by an application to psychological resilience in the Mainz Resilience Project (MARP), which features intermittent longitudinal measurements of stressors and mental health, we propose an approach for individualized, dynamic modeling in this latent space. Specifically, we utilize ordinary differential equations (ODEs) and develop a novel technique for obtaining person-specific ODE parameters even in settings with a rather small number of individuals and observations, incomplete data, and a differing number of observations per individual. This technique allows us to subsequently investigate individual reactions to stimuli, such as the mental health impact of stressors. A potentially large number of baseline characteristics can then be linked to this individual response by regularized regression, e.g., for identifying resilience factors. Thus, our new method provides a way of connecting different kinds of complex longitudinal and baseline measures via individualized, dynamic models. The promising results obtained in the exemplary resilience application indicate that our proposal for dynamic deep learning might also be more generally useful for other application domains.

## Introduction

There currently is a renaissance of artificial intelligence techniques where, e.g., deep learning can provide human-level performance for detecting patterns in images, when trained on large data collections^1^. Deep learning has further been successful with non-image data, where humans cannot directly detect patterns, and also for relatively small sample sizes^2^. Importantly, deep learning has recently been combined with time dynamic techniques such as ordinary differential equations (ODEs)^3–6^, which promises both the advantages of hypothesis-driven modeling and detection of potentially complex patterns. This is why we turned to a combination of deep learning and ODEs when faced with modeling psychological resilience. We propose a novel approach for combining variational autoencoders^7^ (VAEs)—which offer latent representations—with individualized ODEs, i.e., a common system of differential equations where each individual receives an own set of ODE parameters. This approach serves to obtain individualized, anticipated mental health responses to stressor exposure. Empirically, we found imputation and prediction challenging due to the sample size and signal-to-noise ratio. Considering this data situation, our primary goal is to showcase that deep learning approaches can learn individual dynamics even in small-data settings and subsequently identifying associated predictive factors from additional baseline measurements.

Psychological resilience is the maintenance or rapid recovery of a healthy mental state during and after times of adversity^8^. One element of the definition is that both mental health and adversity can change over time and may even do so permanently. Seminal resilience studies^9^ investigated how mental health changes in response to a single potentially-traumatic life event and identified common mental health trajectory patterns such as for resilient individuals (showing stably good or improving mental health in the months or years after the event) and more vulnerable individuals (showing stably poor or worsening mental health). These studies implicitly assume that the observed temporal changes in mental health are due to only *a single stressor event* and that individual differences in the mental health trajectory can be explained by some baseline individual characteristic. However, most individuals are *continuously exposed to more or less severe stressors*. These may include macrostressors (severe life events) but also more “mundane” microstressors, or daily hassles^10^, which are also known to have an impact on mental health^11, 12^. Further, major life events are likely to be followed by other macro- or microstressors^13, 14^. For instance, this means that deteriorations in mental health in the aftermath of a trauma may also have other causes than the trauma. Conversely, not developing long-term mental health problems may be a consequence of only moderate stressor exposure after the trauma. In this case, a resilient trajectory would not result from some protective baseline characteristics but benign life circumstances. The necessity to consider the ongoing, more or less continuous nature of stressor exposure is even more obvious when trying to understand resilience to longer-lasting or chronic adversity, such as adverse life circumstances or life transition phases. In short, investigating resilience should ideally involve repeated longitudinal measurements of stressors and mental health^8, 9, 13, 15^.

This necessity of longitudinal stressor and mental health measurements poses considerable challenges to data acquisition. Usually, mental health and stressor data are acquired intermittently, and not always are measurements equally spaced. The Mainz Resilience Project^12^ (MARP) also faces these problems; self-report measurements are made using online questionnaires every three months *±* two weeks. Additionally, a relevant number of observations (13.7%) are missing and the individual time series differ considerably in the numbers of observations (on average 9.1 observations, with a *min* = 2, *max* = 14, and *sd* = 3.5 in the data set used for learning the individual trajectories). Further, the three-monthly microstressor assessment of daily hassles only covers the past week. In this empirical context—which is prototypical for many longitudinal studies—the challenge is to transform the irregularly sampled mental health and stressor exposure data into continuous time series that are amenable to dynamic analysis.

In the current application, we learn the long-term individual mental health and stressor trajectories in the latent space, obtained from a VAE, with a system of ODEs that allows for encoding assumptions on the interplay of stressors and mental health. The ODE system is parameterized by another neural network, which receives longitudinal summary statistics as input, which also allows for different numbers of potentially irregularly spaced observation time points. On a more abstract level, our proposed new method showcases the versatility of combining deep learning techniques with modeling based on domain knowledge and underlines the usefulness of this particular new method in an application of practical relevance.

A brief sketch of the algorithm is provided next. After this, we show how these individualized models can be linked to baseline measurements for identifying resilience factors, before concluding with a discussion and outlook. In the methods section, we provide more technical details of the proposed method, in particular on how the challenging estimation problem is addressed, and describe the MARP study on psychological resilience, which motivated methods development and is used for illustration. We also provide a Jupyter Notebook on GitLab that shows how our approach can recover structure from exemplary simulated data, and can be used as a starting point for adapting the approach to other applications.

### Related work

Dynamic techniques, such as differential equations, have already been proposed for modeling resilience^16^. Montpetit et al.^17^ presented a coupled version of the multivariate latent differential equation model^18^ for resilience, which estimates the actual values of the latent variable, as well as the slope and curvature simultaneously. Driver and Voelkle^19^ also allow for individualized differential equations. However, both approaches use relatively simple linear transformations for mapping from observed values to latent space. Furthermore, they also require rather strong assumptions on the distribution of differential equation parameters, which may be problematic with respect to the topology in parameter space. In contrast, our proposal strives to combine the advantages of potentially complex non-linear transformations and individualized differential equations, making as little distributional assumptions as possible, as made possible by a combination of VAEs and individualized ODEs.

Numerous deep learning approaches have been developed for longitudinal data; increasingly, they are extended to process data with irregular time intervals, e.g., by using exponential decay functions in the recurrent layers^20–22^. The initial Neural ODE approach^3^ was extended in various directions, also for irregular time series^23, 24^. For enforcing smoothness across time, convolutional inference nets have been proposed, which also can find the lower-dimensional representations and connect them via Gaussian processes^25^. Still others included the temporal information into the VAEs^26–28^ (see Girin et al.^29^ for a comprehensive overview). Yet, neither Gaussian processes nor dynamic VAEs provide an explicit model, i.e., they cannot determine the anticipated stress response. Neural ODEs have been proven successful in learning the dynamics in the latent space in a recent benchmark study^30^ for medical data, which suggests the suitability of ODEs also for our application. To our knowledge, no deep learning approaches were suggested to identify the individualized trajectories in the latent space and continuous time.

Naturally, there are many other techniques—neither based on differential equations nor deep learning—that could potentially be used. For example, growth mixture models identify average starting levels and trajectories and allow for normally distributed deviation from the overall intercept and slope. However, these require an initial dimension reduction step (e.g., calculating a sum score or a factor solution), which is not informed by the longitudinal model and frequently based on rather simple linear transformations. By contrast, our method is designed to solve the problem of dimension reduction and trajectory modeling iteratively. Latent class mixture modeling has also been employed in resilience research^9^ to identify subgroups in the mental health response to trauma, to thereby derive categorical prediction targets (i.e., class membership); we expand on this by providing a set of continuous prediction targets. Psychometric network models have been recently extended to latent variables with autocorrelation^31^. Similarly, dynamic structural equation models^32^ (DSEM) were developed for “intensive” longitudinal data, i.e., data with many observations, and allow for a measurement model and normally-distributed auto-regressive parameters. DSEM also offers analytical tools to investigate the (linear) mappings into the latent space. To our knowledge, neither DSEM nor psychometric network models allow for non-linear mappings into the latent space or joint optimization of the mapping into the latent space and trajectory information. All models in this paragraph, importantly, work by default in discrete time *t* = 1, 2, …, *T*_*i*_ and are not readily applicable to continuous time. In our case, models in discrete time are problematic, considering the missing data patterns and varying observation intervals.

## Results

### An overview of the algorithm, data structure, and training procedure

Figure 1 shows how the data flows through the algorithm. We map observations into a latent space using two separate VAEs^7^ (Figure 1 a). Separating the dimensionality reduction step for mental health and stressor load is a pragmatic decision that introduces some structure onto the learning problem, leaving the modeling of their interaction to the ODE system. The VAEs reduce the number of dimensions to one for each type of measurement (see section ‘MARP data’ below for operationalization). The encoder weights, i.e., the parameters that determine the non-linear mapping, and the decoder weights, which specify the mapping from the latent space back to the level of original measurements, are trained by minimizing the evidence lower bound^7^ based on a Poisson log-likelihood to reflect the count character of the inputs.

**Figure 1.**
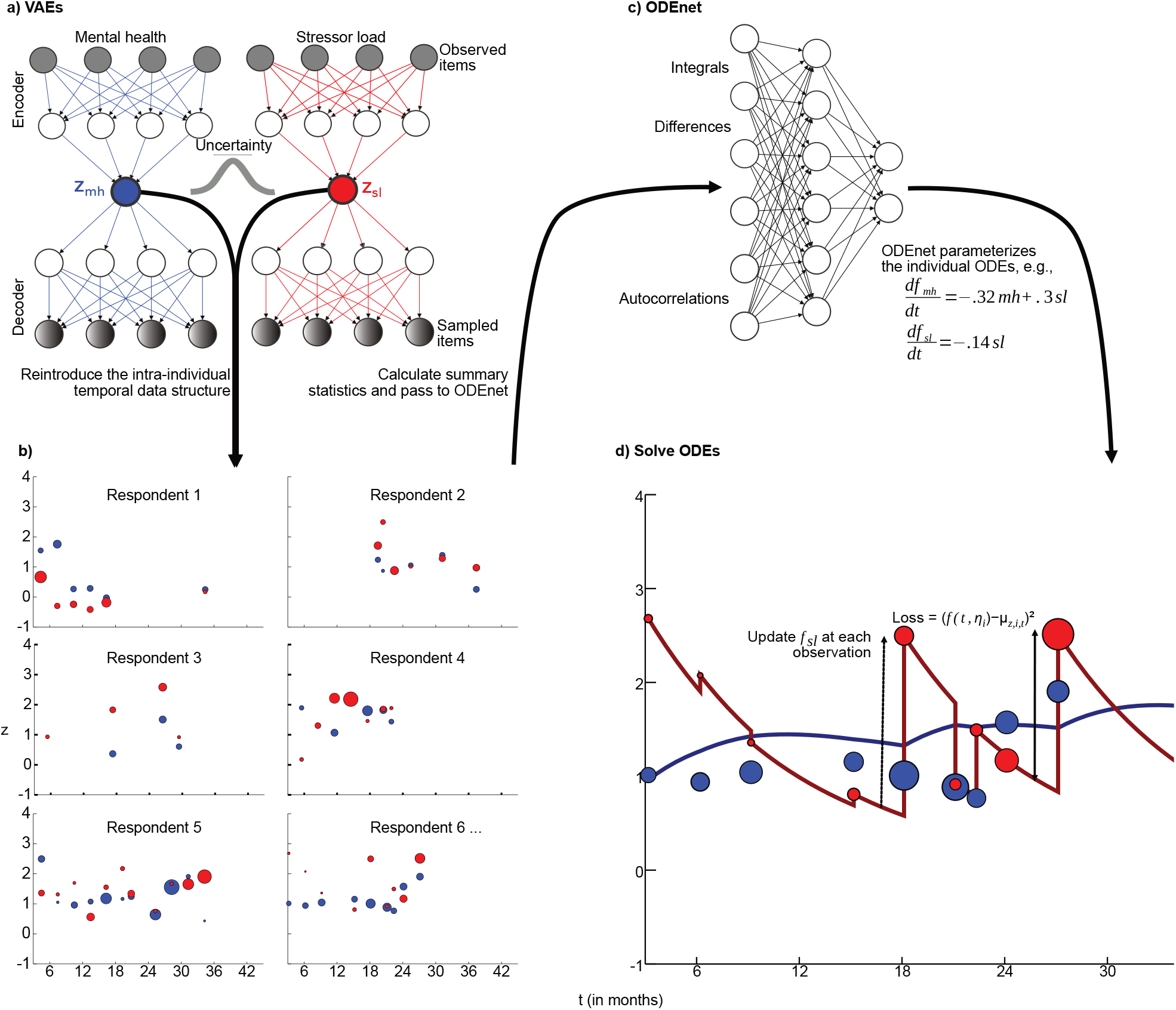
The proposed new method has two essential parts, dimensionality reduction and individualized trajectory estimation. Both tasks are performed using neural networks (upper row). We train two VAEs—one for mental health (blue) and one for stressor load (red)—to estimate the distribution in the latent space for each observation. The variance of these distributions is expressed as size of the dots and reflects uncertainty. Summary statistics of the temporal pattern of latent values are used as inputs to a feed-forward neural network (the “ODEnet”) which is trained to provide ODE parameters that minimize the squared distance of the ODE solution and the latent values, where latent stressor load values are updated at each measurement time point.

The mapping of the observed values in the latent space is performed for all observations with one set of VAE parameters. Afterward, we reimpose the individual and temporal structure, which is essential for further calculations and allows getting first impressions of the latent trajectories (Figure 1 b). Subsequently, each individual time course is aggregated into several summary statistics, again separately for the latent mental health and stressor values. These summary statistics, e.g., include the integral over an interpolated step function and the difference of the first and last values (see Table 1 for a full list), and are chosen such that they do not require fixed observation time points or the same number of measurements for all individuals. These summary statistics then serve as inputs to a feed-forward ODEnet (Figure 1 c), which provides parameters for an ODE system. Thus, optimal combinations and transformations of the summary statistics are determined empirically. For the resilience application, we chose a rather simple ODE system, to reduce complexity when faced with a limited number of individuals and observations. Specifically, the parameters of the ODEnet are trained to minimize the squared distance between the solution of the ODE system and the latent values at the point in time when measurements were obtained from the individuals. We allow the stressor time course to additionally incorporate abrupt external inputs (i.e., sudden changes in the life situation). Accordingly, new observations can override the current state obtained from the ODE. For this purpose, the ODE solver is stopped at the measurement time points, and the latent values for stressors (but not for mental health) are updated. Subsequently, the individualized solutions of the ODE systems are visualized together with the latent values (Figure 1 d). For details beyond what follows, we refer to the online Supplementary Information.

**Table 1.** We include 16 summary statistics—gathered from the individual mappings of mental health (mh) and stressor load (sl) to the latent space (see Figure 1 c)—as inputs to the ODEnet. We additionally scale them according to their range and type (Observation, Difference, Integral, and AutoCorrelation) for achieving numerically similar inputs.

| Input | Type | Scaled by |
| --- | --- | --- |
| First obs of mh | O | 1 |
| First obs of sl | O | 1 |
| First obs mh — last obs mh | D | 1 |
| First obs mh — first obs sl | D | 1 |
| First obs mh — last obs sl | D | 1 |
| First obs sl — last obs sl | D | 1 |
| First obs sl — last obs mh | D | 1 |
| Last obs mh — last obs sl | D | 1 |
| Integral of mh | I | 10 |
| Integral of sl | I | 10 |
| Integral of mh<sup>2</sup> | I | 10 |
| Integral of sl<sup>2</sup> | I | 10 |
| Integral of mh (absolute value) | I | 10 |
| Integral of sl (absolute value) | I | 10 |
| Mean of autocorrelation mh | AC | 100 |
| Mean of autocorrelation sl | AC | 100 |

### ODE system to model resilience trajectories in the latent space

Figure 1 d) shows an exemplary solution of an individualized ODE system, which connects expected values of the distributions in the latent space. We use an ODE system that couples the latent representations of mental health and stressor load. More precisely, we modeled the means of the latent representations depending on the input vectors *µ*(*x*_*i*_) of the latent distributions with the ODE; these *µ*(*x*_*i*_) correlate strongly (*r* ≥ .9) with other methods of dimensionality reduction (see Supplementary Fig. S1 online) and, hence, can be interpreted as valid latent constructs similar to, e.g., more usual sum or factor scores.

The exact design of such an ODE system is a crucial modeling decision since it governs how each component changes and, accordingly, requires domain expertise. Given our sample size and sampling frequency, we work with a rather simple ODE system to keep the number of estimated ODE parameters *η*_*i*_ small. Specifically, we use the system

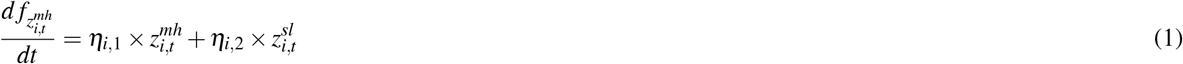

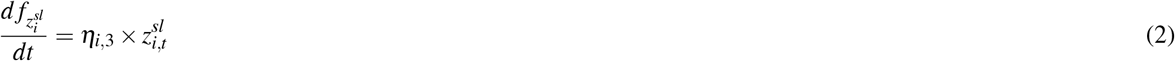

where changes in 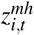, the latent value reflecting mental health problems, and 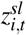, the latent value reflecting stressors load, are driven by the current own value. Additionally, mental health changes in response to stressor load. Separate parameter values *η*_*i*,1_ − *η*_*i*,3_ for each individual *i* allow for individualized trajectories of mental health and stressors.

Negative values for *η*_*i*,1_ and *η*_*i*,3_ effectively realize system-inherent damping^16^, where high mental health and stressor values, respectively, are more quickly driven back to low latent mental health and stressor values. Thus, a high negative *η*_*i*,1_ in particular reflects good recovery from mental health problems (since the majority of observations are mapped into the positive valued latent space or close to zero, see Figure 1 c). Positive values for *η*_*i*,2_ realize the adverse effect of stressors on mental health. A low positive *η*_*i*,2_ value thus reflects a low mental health impact of stressors in the individual.

At each realized measurement, the value of the integrator of the latent stressor load 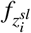 is updated to the mapping of the actually observed value 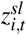. This reflects the obvious notion that stressor levels are only partly driven by an endogenous property of the ODE system (i.e., damping) but mainly reflect exogenous forces, that is, the sudden occurrence or absence of stressors that lead to abrupt changes in the latent stressor values (see above).

The benefits of such an ODE system in comparison to discrete-time models like regression are crucial for analyzing the data from the MARP study at hand. Most importantly, differential equations take all available information at the precise time into account. Thereby, irregular sampling intervals and entirely (or partly) missing observations are dealt with by the properties of our dynamical system (assuming non-informative missingness).

### Person-specific ODE parameters

The ODE parameters for every respondent are estimated in an unrestricted manner, i.e., without explicit constraints regarding their distribution or direction. This approach allows for widely different dynamics, which we refer to as individual dynamic models while the models’ overall structure stays stable. This is accomplished by estimating the parameters of a fixed ODE system *η*_*i*,1_ to *η*_*i*,3_ with the ODEnet—a feed-forward neural network—with 16 summary statistics as inputs (see Figure 1 c and Table 1 for details). While we are using linear ODE system for the specific resilience application, i.e., also an analytical solution exists, the proposed approaches enables more complicated non-linear systems. Also, the non-linear mapping to the latent space via the VAEs may pick up some non-linearity in the trajectories, helping to model the latent dynamics with this reasonably simple ODE system adequately. Importantly, the summary statistics are selected with data availability in mind. Since some MARP participants just started recently to provide data, all summary statistics need to be computable with only two observations of mental health and stressors (not necessarily at the same time), which is the minimum requirement to be included in the longitudinal analysis.

The ODEnet provides the individual parameters of the ODE system as outputs. Accordingly, the overall structure of how mental health and stressors change is the same for all respondents and specified by the ODE system. Yet, the values of the summary statistics—included in the ODEnet—differ from person to person. Therefore, each individual receives an own variant of the ODE system. The ODEnet is trained to minimize the squared difference between the ODE solutions 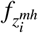 and 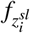 and the latent values 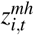 and 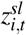, using a standard neural network backpropagation algorithm. In contrast to previous methods (e.g., growth mixture models), the individual parameters do not obey any predefined distribution; they can be estimated in the absence of such restrictions by optimizing the fit to the data.

### Quantification and prediction of resilience

In addition to longitudinal measurements of mental health and stressor load, the MARP study incorporates a large battery of additional baseline measurements for identifying potential resilience factors, i.e., characteristics of individuals that might predict reduced reactivity of mental health to stressor load. For identifying such resilience factors, we can either predict the individual parameters *η*_*i*_ directly or apply an artificial stress test to each individual (see Figure 2). This artificial stress test means that the latent mental health value of an individual is initialized to an average value, and the latent stressor load value is set to an initial value that corresponds to a high stressor level. The stress reaction pattern of each individual then is characterized by how the latent values develop according to the individualized ODE system, if no further external input is provided. Predicting each of the ODE parameters separately allows for investigating the different dimensions of resilience in our model (i.e., recovery of mental health (*η*_*i*,1_) and reactivity of mental health to stressors (*η*_*i*,2_)) independently. In contrast, the artificial stress test considers all parameters jointly, to obtain a holistic picture of resilience. This also serves as a diagnostic tool to investigate whether the training of the algorithm succeeded. We choose four points in time (after 5, 9, 15, and 20 months) and predict the individual mental health values with measurements from the baseline battery, covering proteomics, behavioral measures, and questionnaires.

**Figure 2.**
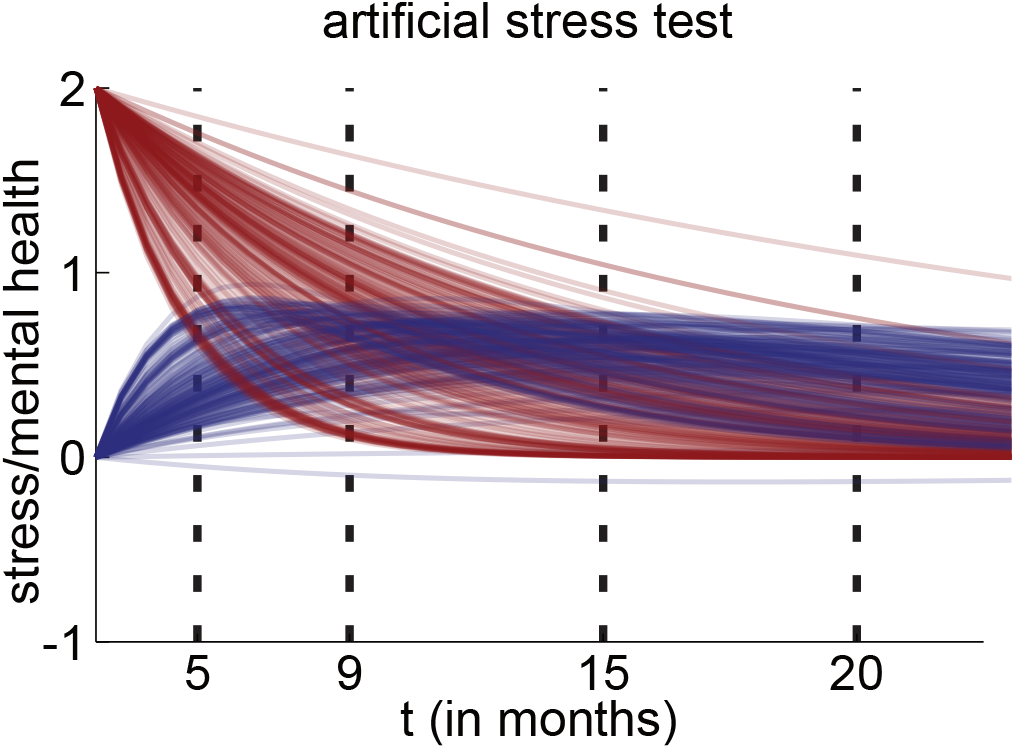
The artificial stress test induces a considerable amount of stress 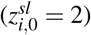 on each individualized ODE system and captures how the mental health (blue) and stressor load (red) are predicted to develop in the latent space (y-axis) over time (x-axis). Prediction targets are derived from the values of mental health at predefined time points (ptp 1-4) of 5, 9, 15 and 20 months.

Specifically, we use the lasso, a regularized regression approach that provides variable selection^33^. The amount of regularization is chosen according to prediction performance, specifically by 6-fold cross-validation. For a robust result, this procedure is repeated 1000 times on resampling data sets, obtained by sampling with replacement, resulting in inclusion frequencies^34^. Thus, a baseline measurement would receive inclusion frequency equal to or close to zero, if there would be no connection between the baseline measurement and our proposed quantification of resilience by the stress test. Quite in contrast, we find rather strong relations between our derived quantification of resilience and some baseline measurements, which may thus be considered to be potential resilience factors (see Figure 3). Since the present paper has its focus on the proposed new method, and not on resilience research, we do not report the selected variables. We compare the prediction targets derived with the artificial stress test to those of an auto-regressive random effects model (see online Supplementary Figure S2) with other latent variable models. The number of prediction targets of our approach is comparable to the most successful competitor. We find some overlap between the different approaches, which indicates that our approach serves as a complement to conventional methods to quantify individual differences.

**Figure 3.**
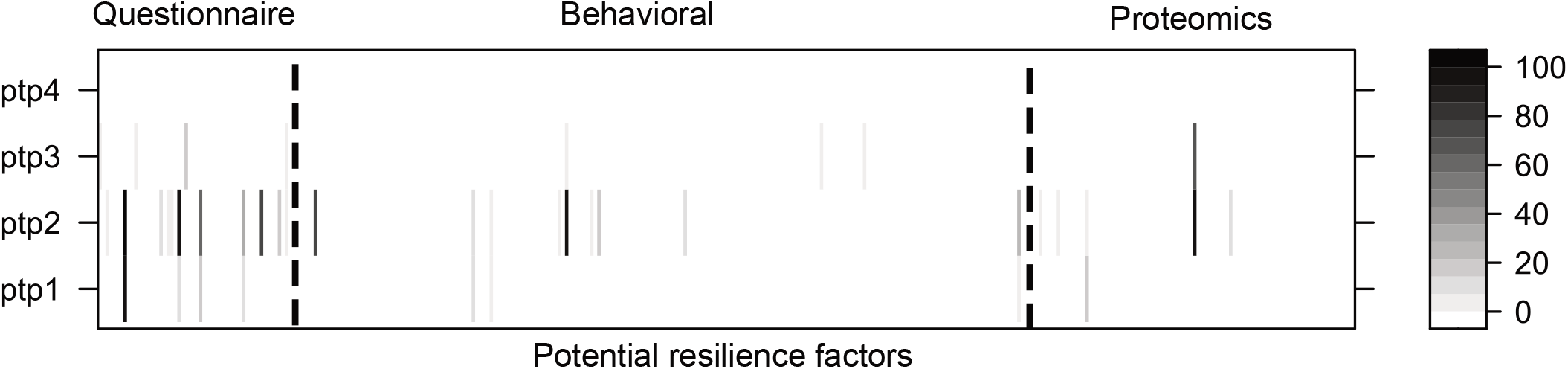
Inclusion frequencies of potential resilience factors in % (shades of gray) using a cross-validated lasso analysis (*n* = 88; *p* = 350; 1000 repetitions) predicting mental health at predefined time points (ptp) of the artificial stress test, i.e., the intersection of the blue lines with the vertical, dotted lines in Figure 2). We identified potential resilience factors in all three data modalities (i.e., questionnaires, behavioral measures, and proteomics).

## Discussion

We combine deep generative learning with a system of ODEs to obtain individual parameters of a dynamic model, and illustrate this novel approach in an application on psychological resilience. The proposed method can handle problems typical for real-world applications, specifically irregular measurement time points and limited sample size. In particular, the augmentation of neural networks with ODEs allows us to incorporate domain expertise, which can dramatically reduce the sample size requirements of deep learning^6^. An important ingredient of this approach is differential programming^35, 36^, i.e., automatic differentiation in complex models, which allows for tailoring algorithms to particular needs, such as combining neural networks with ODEs. This framework is more specifically used in our proposed method to allow the integrator of the ODE solver to jump to updates of the latent stressor load values.

We show that this flexible, non-linear, and versatile combination of scientific modeling and machine learning can be utilized to quantify resilience and identify resilience factors, and agree with others^3, 6^ that neural differential equations promise considerable opportunities for this particular field and other disciplines working with longitudinal data.

From a subject matter point of view, the most substantial benefit of our approach are the resulting individual parameters of the dynamic model. The approach builds upon the overall structure of stress research, according to which mental health problems change due to stressor load, and incorporate the main lesson from resilience research that there are significant individual differences in how people deal with stressors and recover after stressful episodes. In fact, these individual differences are what resilience research is investigating. Instead of reporting population-level prevalence (like, e.g., what percentage of people can be labeled as resilient) we are interested in modeling the mental health trajectory of each person in response to his or her stressor load. This perspective on each individual also addresses the needs of health professionals who are interested in personalized predictions tailored to their particular client, strongly informed by his or her unique history. Hence, we are interested in the most flexible approach that allows for capturing personal nuances best. Compared to this, we argue that it is of secondary interest (at most) to assign people to categorical labels like resilient and vulnerable or force individual deviations from the grand mean to be aligned with predefined distributions. In this regard, we argue that the continuous individual differences—expressed here as ODE parameters and harnessed with an artificial stress test—are meaningful quantifications of resilience and help to identify resilience factors from large baseline batteries.

A limitation of the proposed model is the simplicity of the ODE system. There are valid theoretical reasons to assume that resilience might be better captured using an additional second-order derivative (i.e., acceleration) of mental health problems. While our first-order ODE system was a deliberative decision in consideration of the small number of observations and individuals (for dynamic modeling and deep learning standards at least), future research might experiment with more complicated, e.g., non-linear systems of differential equations which we found hard to train with this approach and data set. Due to our sample size, we also equate the expected value *λ* with the dispersion and assume a Poisson distribution in the generative part of both VAEs. Loosening this admittedly strong assumption would require us to learn—at least in the VAE setting—thousands of additional parameters, which we refrained from. The current application, however, focuses predominantly on the trajectories in the latent space. Accordingly, we expect little benefits from learning the dispersion in the decoder network. Also, future work might investigate the prediction and imputation capabilities of this approach and compare it to existing methods^20, 25, 37, 38^. Another interesting direction for future research is to soften the distinction between mapping the observations in the latent space and learning the trajectories with ODEs; e.g., by including trajectory information into generative learning (as dynamic VAEs^29^ do) Also, our approach assumes stationarity and accordingly neglects the possibility of long-term learning and plasticity (i.e., temporal changes in resilience). While we could extend our model to weaken this assumption, our main focus in this work is on the identification of resilience factors based on all available longitudinal measures. While we concentrated on learning the individual dynamics, future work might extend this by learning the individual equilibria. Furthermore, we base our hyperparameter choice mainly on sample size requirements and conventions. There are, however, more principled ways to determine these decisions, especially in the realm of AutoML^39^. However, such strategies usually require data splitting, which we do not apply to tune our deep learning components due to the relatively small sample size. We improved interpretability compared to purely black-box approaches due to the specifications in our ODE system; however, we lack the interpretability of combinations of classical approaches, e.g., an ARMA model based upon confirmatory factor analysis. We argue, however, that we sacrificed some interpretability to gain considerable flexibility regarding, e.g., the non-linear mapping of our VAEs and the jump equation of our dynamical system. Future work will investigate repeated baseline observations and time-dependent predictors. Yet, already the individualized dynamic models enabled by our new method provide an important building block for better understanding psychological resilience. Therefore, we are confident that other applications that require individualized parameters of dynamic models may benefit from our approach.

## Methods

### Dimensionality reduction per time point with VAEs for count data

We choose variational autoencoders (VAEs) to find a lower-dimensional latent representation because they provide a flexible framework with numerous potential extensions and applications. VAEs comprise a recognition model and a generative model. The purpose of training the recognition model (aka inference model or encoder) is to find the variational parameters *φ* of a neural network to approximate the posterior distribution of the latent variable *z* given the inputs *x*, i.e., *q*_*φ*_ (*z* | *x*). The parameters of the generative model (aka decoder) *θ* are trained to increase the log-likelihood of the inputs given random samples from the posterior distributions. To train the recognition and generative model simultaneously, we maximize the evidence lower bound (ELBO) of the marginal log-likelihood

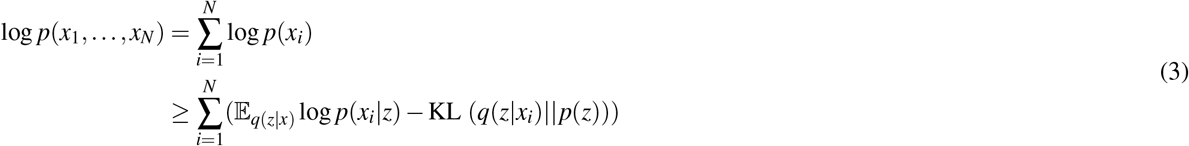

where the first term of the right hand side is the expectation of the log-likelihood of *x*_*i*_ given *z* with respect to *q*(*z* | *x*). The Kullback-Leibler divergence (*D*_*KL*_) penalizes deviations of the posterior from the prior. Temporal dependence between the observations is established by the system of ODEs. For computation, we plug in the Poisson log-likelihood and the closed form of *D*_*KL*_ for a Gaussian prior and posterior. Thereby, our training objective becomes

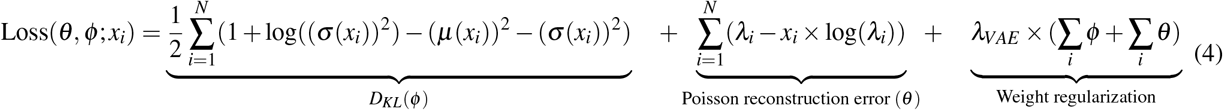

where *µ* and *σ* are the mean and standard deviation of the latent distribution depending on the observed values *x*_*i*_. *λ*_*i*_ is the expected value of the Poisson distribution of each respondent and item. To prevent overfitting, we regularized the encoder (*φ*) and decoder (*θ*) weights of both VAEs with *λ*_*VAE*_. The Poisson distribution does not perfectly fit the data since we found moderate levels of overdispersion for some of our items. Yet, the Poisson distribution avoids an additional parameter, which might be difficult to determine, by coupling expected value and dispersion. We used *J* nodes and *tanh* activation functions in the middle layers. In the final layer, we used a *ReLU* activation function to strictly pass non-negative values to *λ*. All neural networks were trained with Flux.jl^40^ and the Adam^41^ optimizer (see Training section below and online Supplementary Information for details). We used this website^42^ as a basis for drawing the VAEs (in Figure 1 a) and ODEnet (in Figure 1 c).

### Individual ODE parameter estimation with the ODEnet

The parameters of the ODEnet *τ* were trained to minimize the sum of the squared difference of the trajectory 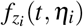 and the mean of the latent space distribution 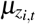 at the precise point in time *t*. Accordingly, the loss function for the ODEnet is

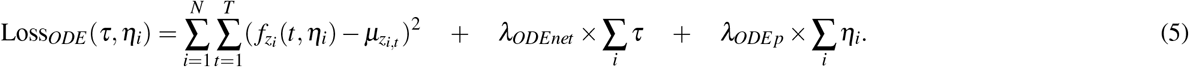

The ODEnet is a separate feed-forward neural network with two layers and 16 inputs; the mid-layer and end-layer have three nodes (due to data size considerations). As inputs, we use a mixture of integrals, first and last observations, differences, and autocorrelations (see Table 1).

We used a piecewise constant step function to approximate the integral of the individual trajectories defined as 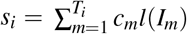 where *c*_*m*_ is the value of 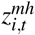 or 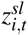 on the interval *I*_*m*_ = (*t*_*i*−1_, *t*_*i*_) and *l*(*I*_*m*_) the length (*t*_*i*_−*t*_*i*−1_) of this interval. We also included 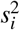. It was left to gradient descent to find a good combination of these inputs to minimize Loss_*ODE*_ (*τ, η*_*i*_) given the value of the ODE at a particular point in time 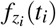 which, in turn, depends on the individual set of ODE parameters *η*_*i*_.

We use a *ReLU* activation function in the middle layer and no transformation in the final layer. The choice of inputs is restricted by the minimum requirement of two observed values for each group of variables at any point in time. We scale the inputs to ensure approximately equal numerical size. For reasons explained above, we update the integrator at every realized measure of 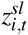. Such steps are implemented in DifferentialEquations.jl^43^ and differentiable through neural nets via DiffEqFlux.jl^44^. All solved ODE systems start the first measurement 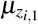. To deal with unit non-response, Loss(*θ, φ*; *x, λ*_*VAE*_) and Loss_*ODE*_ (*τ, η*_*i*_) are only evaluated at actual measurement time points.

### Training

Experimenting with different training strategies, we obtained a sufficiently low

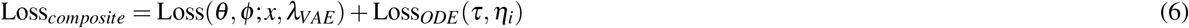

with a combination of separate pretraining of the VAE and ODEnet combined with a sequantial training of both. More detailed, we pretrain both VAEs with a learning rate *α* = 5^−5^ for 80 epochs. Then, we pretrain the ODEnet for 50 epochs with *α* = 1^−6^. Afterward, we sequentially train the VAEs and the ODEnet in the same epoch with *α* = 1^−6^ for 100 epochs. We also compared this approach to a joint training of all involved components. We find the sequential training strategy to work better for this purpose. We did not systematically tune our hyperparameter choices (see Supplementary Table S1). While more principled^39^ ways to determine hyperparameter choices would have led most likely to a lower Loss_*composite*_, this approach avoids heavy overfitting. All neural networks were trained with the Adam optimizer^41^ and standard decay rates (*β*_1_ = 0.9 and *β*_2_ = 0.999).

### MARP Data

The Mainz Resilience Project (MARP) is an ongoing study that started in 2016 with a planned study duration per participant of seven years and is conducted by the University Medical Center Mainz and the Leibniz Institute for Resilience Research^12^. All available observations (*N*_*sl*_ = 1.233 and *N*_*mh*_ = 1.196) were used to train the VAEs. Each observation consists of 28 mental health items (GHQ-28^45^) and 58 daily hassle (MIMIS battery^46^) items. One item is a question to either rate 28 different health aspects on a scale from 0-3 (mh) or indicate the days last week (0-7) where certain unpleasant circumstances were experienced (sl). This vector of length 28/58 was plugged into the respective VAE and mapped into the latent dimension. To be included in the longitudinal analysis, at least two observations of mental health and stressor load—at any point in time within the observation window of 3.5 years—were necessary. This basic requirement was met by *N* = 166 participants. Four respondents were excluded due to unusual patterns. In this longitudinal data set, on average 9.1 observations (*min* = 2, *max* = 14, *sd* = 3.5) per person are available. The majority of the longitudinal measures are gathered via an online assessment roughly every three months. Calendar time was protocolled at logging in to the online questionnaire. Study time was taken by subtracting the calendar time of each observation with the first baseline observation. Accordingly, observations of stressor load and mental health got the same study time, although they were not taken simultaneously but during the same login. Besides the request to provide data on their mental health and stressors (daily hassles and life events, of which only daily hassles are used here), the participants went through an extensive baseline examination, which includes in total *p* = 350 neuroimaging, behavioral, proteomics, and survey data variables. One hundred fifty-five participants met the longitudinal requirements; hence, they underwent the artificial stress test (see Figure 2). However, only *N*_*lasso*_ = 88 respondents had a complete baseline battery and sufficient longitudinal information for being included into the lasso analysis. Participants were recruited in a critical life phase aged 18 to 20 at study inclusion with a prehistory of critical life events.

## Data Availability

Data cannot be made publically available at the moment.

## Acknowledgements

This project has received funding from the European Union’s Horizon 2020 research and innovation programme under grant agreement No 777084 (G.K., S.P., H.E., O.T., R.K.) and the State of Rhineland-Palatinate, Germany (MARP program, DRZ program, Leibniz Institute for Resilience Research; M.K., A.Sch., M.W., O.T., R.K.).

## Author contributions statement

H.B. and G.K. developed the idea, constructed and tested the models, and wrote the paper. All authors contributed through discussions and reviewed and approved the manuscript.

## Additional information

### Human participants

All methods were carried out in accordance with relevant guidelines and regulations. All experimental protocols were approved by ethics committee of the Medical Board of the State of Rhineland-Palatinate, Mainz. All subjects gave informed consent.

