## Supplementary material for "Individualizing deep dynamic models for psychological resilience data": online Supplementary Information

### Introduction

This file supplements the findings of the paper. More precisely, it compares different ways to reduce the dimensionalities, introduces other ways of quantifying resilience and investigates their predictability, investigates whether more restrictions on the parameter space benefit the algorithm’s training, and provides details on the parameter choices.

### Comparison of the VAE $\boldsymbol{\mu}$ to other dimensionality reduction techniques

Applying VAEs for dimensionality reduction invites criticism concerning the validity of the lower-dimensional representations. This section compares the $\mu$ of the VAE to more conventional ways to reduce the number of dimensions. More detailed, it tests the assumption that the VAE $\mu$ can be interpreted similarly to more transparent ways to find lower-dimensional representations.

A straightforward way to find lower dimensional representations is to use the sum score, i.e., sum all items up (as they all share the same direction). One might also apply principal components analysis to find uncorrelated representations of the scales, or with only one dimension to find a linear transformation of the data with the largest variance. A third method—used especially by psychometrically trained people—is the confirmatory factor analysis. Due to its specialized origin, an analyst can choose to base their dimensionality reduction on the product-moment correlation for continuous variables or polychoric correlation tailored to Likert-item scales with a low number of answering possibilities (e.g., four as in the GHQ scale).

Please note, the PCA approach makes less sense for the stressor load scale than for the mental health scale since the stressor load is not meant to identify one (or more) uncorrelated latent dimensions; instead, the item battery of stressor load aims to capture most of the daily hassles people may experience. The primary aim of this is analysis is, however, to compare different dimensionality reduction techniques also in applications similar to the one shown in the paper. For this reason, the subject matter problem of no intended single latent construct is ignored in what follows.

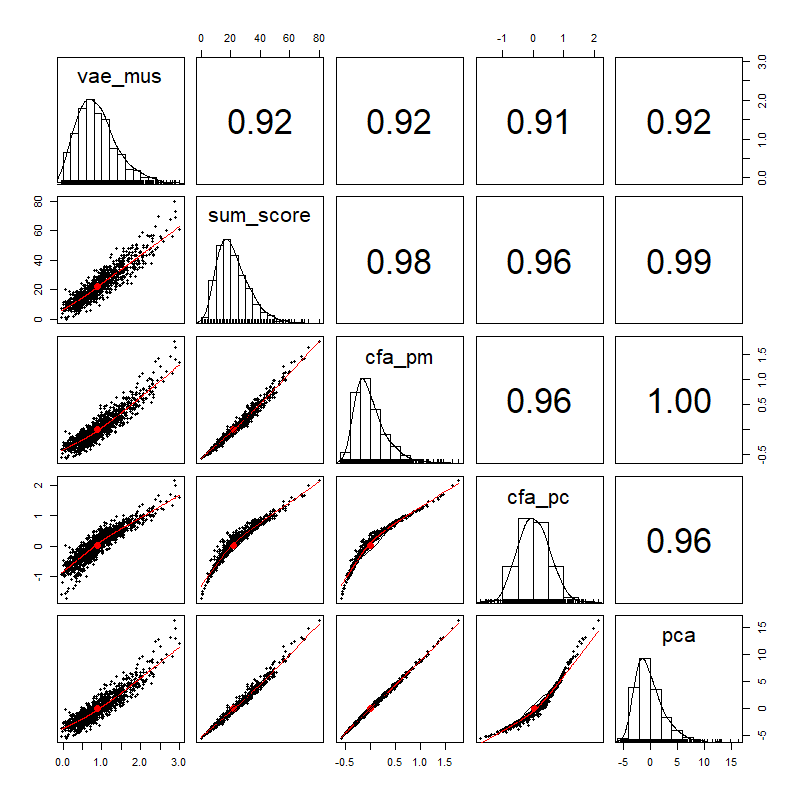

Supplementary Fig. S1: Scatterplot matrix investigating the differences between several dimensionality reduction techniques.

While the correlations (see upper diagonal of Figure S1) between the VAE and the others are the weakest, they are all $>.9$, indicating a high agreement between the dimensionality reduction techniques. Also, the correlation between the CFA based on product-moment and polychoric correlations is very high. This high correlation indicates that potential refinements concerning the measurement scale do not endanger the fundamental conclusions drawn in this paper.

### Comparison to other methods of quantifying resilience

There are numerous ways to quantify resilience; only one of them is the NODE-VAE enriched with the artificial stress test. Another intuitive way to quantify resilience is to estimate random effects models of individual mental health ($mh_{t,i}$) with lagged mental health ($mh_{t-1,i}$) and stressor load ($sl_{t-1,i}$). In this model, the random slopes of mental health but particularly stressors provide information on how far and in what direction respondents deviate from the average stress response. More detailed, the model has the following regression equation

$$mh_{t,i}=\beta_{0i}+\beta_{1i}\times mh_{t-1,i}+\beta_{2i}\times sl_{t-1,i}+\epsilon_{i}$$

with $\beta_{0i}=\gamma_{00}+u_{0i}$ as random intercept, $\beta_{1i}=\gamma_{10}+u_{1i}$ , and $\beta_{2}i=\gamma_{20}+u_{2i}$ as random slopes. Importantly, the individual variability $u_{1i}$ and $u_{2i}$ assumed to be distributed normally with $u_{0i}\sim N(0,\sigma_{uoi}^{2}),u_{1i}\sim N(0,\sigma_{uoi}^{2})\text{, and}u_{2i}\sim N(0,\sigma_{uoi}^{2})$ around the main effects $\gamma_{10}$ and $\gamma_{20}$ provide a additional prediction targets as, e.g., Snjiders and Boskers point out in Chapter 5.2.

Next to the way to quantify individual differences, we also emphasized the different ways to arrive at the latent representations. Accordingly, a natural question is whether this quantification can benefit from altering the dimensionality reduction techniques, measured as differential predictability by the potential resilience factors. Accordingly, Figure S2 investigates the predictability of the different components, i.e., auto-regression and the strength of the previous stressors on health.

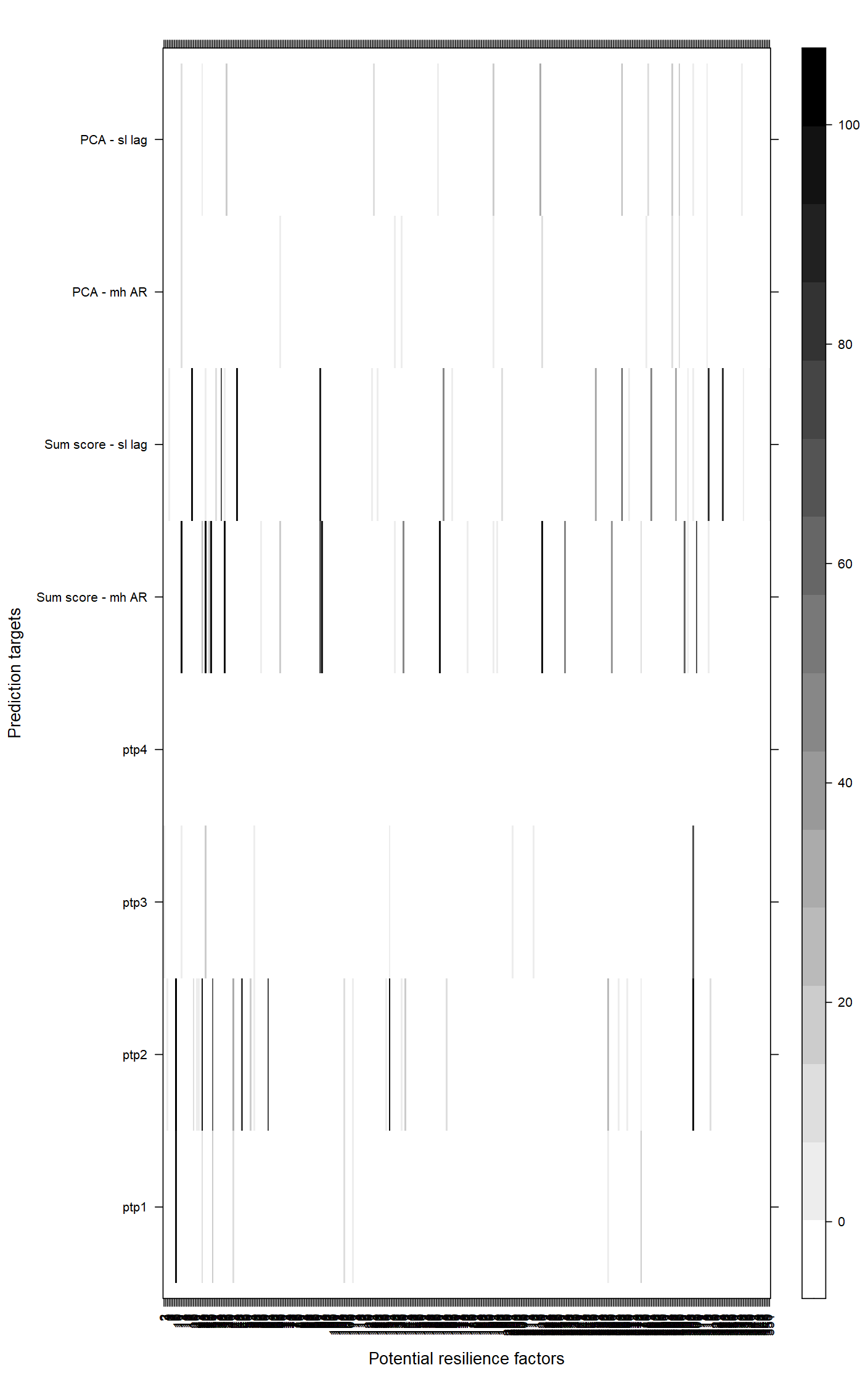

Supplementary Fig. S2: Inclusion frequencies of auto-regressive random effects in comparison to the artificial stress test.

Figure S2 clearly shows that the PCA approach combined with the random effects models provides a limited number of potential prediction targets, which tend to be rather unstably selected by the cross-validated lasso. In contrast, the sum score approach provides 74 potential prediction targets. However, the majority of 48 potential prediction targets are identified for the auto-regressive term. We argue that this is a meaningful metric for health stability; however, it tells us little about the respondent’s stressor reactivity. In contrast, the predefined time points (ptp1-4) investigate how respondents react to a considerable amount of stress. Accordingly, this metric is tailored to our primary interest in health trajectories in response to adverse circumstances.

There is limited accordance between the different procedures. Some hits were close to each other, indicating that they often were found in the same questionnaire battery or task but different subscales or extractions. Accordingly, the artificial stress test acts as a complement to conventional approaches.

### Constraining the parameter space of the ODEs

We allowed the ODEnet to insert unconstrained outputs into the system of differential equations. This freedom follows our line of argumentation that we do not want to impose any assumptions on the range and shape of the parameter estimation (in contrast to the ODE system where we deliberately deviate from this position). A reasonable criticism of this approach is that assumptions on the ODE parameter space might increase this algorithm’s efficiency.

To investigate this potential line of criticism, we ran the following experiment. We trained 90 ODEnets with different initializations and the following three activation functions:

$$\sigma_{1}(x)=x$$

$$\sigma_{2}(x)=(1\div(1+\text{exp}(-x)))-.5$$

$$\sigma_{3}(x)=(1\div(1+\text{exp}(-x)))-.5)\times2$$

where $\sigma_{1}(x)$is the identity function, $\sigma_{2}(x)$ is the sigmoid function $-.5$ to center the parameter space around $0$, and $\sigma_{3}(x)$ multiplies $\sigma_{2}(x)$ with 2 to provide a larger possible parameter space.

In every run, we identified the minimum sum of all deviations between the ODE trajectories $f_{z_{i}^{mh}}$ and the actually observed values $z_{i}^{mh}$ of mental health. Figure S3 shows the minimal sum of distances for the different seeds.

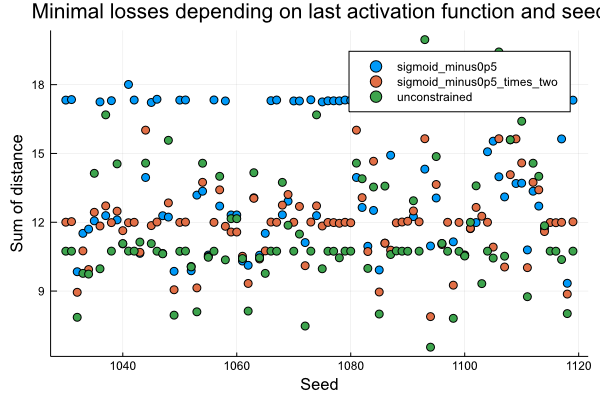

Supplementary Fig. S3: Minimal sum of distances from health trajectory to observation (mu) with the same 90 ascending seeds. Unconstrained output shows the smallest deviation but also the largest distribution.

The lowest overall sum and median of distance is achieved by the unconstrained training ($\sigma_{1}$, green, median $=11.6$) in contrast to $\sigma_{2}$ (orange, median = 12.0) and $\sigma_{3}$ (blue, median = 13.35). At the same time, however, the unconstrained input into the ODEnet also achieves the largest sum of distance. Accordingly, one might conclude that unconstrained training leads to the best, i.e., closest fits between the latent variables and their distributions; however, it also requires more care when training the model to arrive at those close-fitting solutions.

### Parameter choices

Among the practical considerations of every tailored, deep learning approach are the choices of the hyperparameters. Table S1 provides an overview of the main hyperparameter choices. We restricted the number of latent variables to the bare minimum of one per group of items, i.e., mental health or stressor load. Next to the ease of interpretation, the main reason for this restriction is that more ODE parameters increase training difficulty given the number of data points.

Supplementary Table. S1: Justification of hyperparameter choices

| Parameter type | Parameter | Justification |
| --- | --- | --- |
| Number of hidden layers | 1 | Sample size restriction |
| Number of nodes in hidden layers | 58 (sl VAE), 28 (mh VAE), 3 (ODEnet) | Size of the input or output |
| Latent variables | 1 | Interpretability, difficulties to train ODEnet providing many ODE parameters with a small number of respondents |

The number of hidden layers was also determined with the sample size in mind. Since the number of observations and respondents is low for deep learning and dynamic modeling standards, we restricted this number also to its minimum. The number of nodes in the hidden layers follows the simple heuristic that it should equal the number of inputs for the VAE decoder and the number of outputs for the VAE encoder and the ODEnet.
